# Temporal deep learning with clinically engineered biomarkers for the early prediction of type 2 diabetes

**DOI:** 10.1101/2025.11.26.25341040

**Authors:** Iqra Naveed, Mohammad Noaeen, Mohammed A. AboArab, Muhammad Farhat Kaleem, Karim Keshavjee, Aziz Guergachi

## Abstract

Diabetes mellitus remains a major global health burden, causing an estimated 3.4 million deaths in 2024 and highlighting the need for accurate early identification of individuals at risk of developing type 2 diabetes (T2D). Electronic health records (EHRs) provide longitudinal clinical trajectories, yet many predictive frameworks fail to capture short-, intermediate-, and long-term temporal patterns or incorporate clinically validated metabolic biomarkers. This study introduces a hybrid deep learning framework that integrates hierarchical temporal modeling with clinically engineered predictors for early T2D risk estimation. The approach includes data preprocessing, temporal sequencing, and the incorporation of derived biomarkers such as triglyceride-to–high-density lipoprotein cholesterol ratio (TG/HDL-C), low-density lipoprotein to high-density lipoprotein cholesterol ratio (LDL/HDL-C), total cholesterol to high-density lipoprotein cholesterol ratio (TC/HDL-C), very low-density lipoprotein (VLDL), obesity status, and prediabetes indicators. A multilevel convolutional neural network (CNN) extracts low-, mid-, and high-level temporal features, which are processed in parallel by long short-term memory (LSTM) modules to capture multi-scale temporal dependencies. The fused temporal and biochemical representations form a unified CNN–LSTM architecture that is evaluated using standard classification metrics. Experiments conducted on 19,218 patients and 368,790 clinical visits from the Canadian Primary Care Sentinel Surveillance Network (CPCSSN) achieved 93.2% accuracy, 75.7% sensitivity, 98.8% specificity, and an 84.4% F1 score, outperforming bidirectional long short-term memory (Bi-LSTM), support vector machine (SVM), k-nearest neighbor (KNN), and baseline CNN–LSTM models. Feature importance analysis identified fasting blood sugar (FBS), glycated hemoglobin (HbA1c), and lipid ratios as the strongest predictors. By combining temporal representation learning with clinically grounded biomarkers, the proposed framework provides an interpretable, scalable, and robust foundation for early diabetes risk prediction and can be extended to other chronic diseases characterized by longitudinal EHR data.

**Author Summary:** In this study, we focus on the growing challenge of type 2 diabetes, a condition that develops gradually and often remains undetected until significant health damage has occurred. Our goal was to create an approach that identifies individuals at increased risk much earlier by examining how their clinical measurements change over time. To achieve this, we analyzed routine health information collected during repeated medical visits and combined it with key biological markers known to reflect metabolic health, such as blood sugar levels, long-term glucose measures, and cholesterol-related indicators. We developed a computational model that learns how these factors evolve and how they relate to the future onset of diabetes. When tested on a large population dataset, our model detected risk patterns more accurately than several widely used prediction methods. We also found that variations in blood sugar, long-term glucose, and lipid measures played a particularly important role in identifying individuals likely to develop the disease. By offering earlier and more reliable risk assessment, our work supports more proactive and personalized preventive care. Ultimately, this approach has the potential to help clinicians intervene sooner and reduce the burden of diabetes-related complications.

## I. Introduction

Type 2 diabetes (T2D) is one of the most common chronic metabolic diseases. It causes persistent high blood sugar due to impaired insulin secretion, reduced insulin sensitivity, or both [1, 2]. This disease can lead to serious complications that affect the eyes, kidneys, nerves, and cardiovascular system. The global burden is increasing at a rapid pace. The International Diabetes Federation (IDF) estimates that 589–590 million adults were living with diabetes from 2024–2025. This represents approximately one in nine adults, and more than 40% of them are not diagnosed [3, 4]. Mortality is also increasing. Diabetes caused an estimated 3.4 million deaths in 2024, which equals one death every nine seconds. According to the World Health Organization (WHO), 1.6 million deaths were directly linked to diabetes in 2021, with almost half occurring before the age of 70 [1]. The financial cost is high. Global diabetes health spending reached US$1.015 trillion in 2024, which accounts for almost 12% of total health expenditure [5]. Early detection of people at high risk of T2D can prevent or delay complications. Clinical guidelines recommend structured risk assessment and intervention [6, 7]. Traditional risk models, such as the QDiabetes model, use demographic, clinical, and laboratory features to estimate long-term risk. These models are useful at the population level but are limited. They rely on cross-sectional data and do not reflect the dynamic course of the disease. Electronic health records (EHRs) provide longitudinal information that shows how T2D develops over time. This makes them a valuable resource for risk prediction.

Recent studies have shown that machine learning and deep learning models improve prediction accuracy when applied to EHR data [8-10]. These methods can detect patterns in patient records that are not captured by conventional models. However, important gaps remain. Many studies ignore short- and medium-term trends that appear before the onset of the disease. Others fail to include clinical markers that reflect the underlying metabolic state. Lipid ratios and obesity indicators are among the strongest predictors of T2D. The triglyceride-to–high-density lipoprotein cholesterol (TG/HDL-C) ratio is strongly associated with incident T2D and performs better than many standard lipid measures do [11]. The low-density lipoprotein to high-density lipoprotein cholesterol (LDL/HDL-C) ratio and the total cholesterol to high-density lipoprotein cholesterol (TC/HDL-C) ratio, as well as obesity status, also reflect insulin resistance and metabolic imbalance. The inclusion of these features in prediction models can enhance their performance and clinical value.

This study presents a hybrid deep learning framework that combines temporal features with clinically engineered biomarkers for early T2D risk prediction. The framework extracts low-, mid-, and high-level temporal features through convolutional pooling layers. Each level is then processed in a separate recurrent module. The outputs are fused with clinical indicators such as lipid ratios and obesity measures. This study makes several key contributions. (i) The proposed framework introduces a multilevel temporal modeling strategy that preserves detailed disease progression patterns. (ii) It integrates TG/HDL-C, LDL/HDL-C, TC/HDL-C, very low-density lipoprotein (VLDL), obesity, and prediabetes status to improve interpretability and accuracy. (iii) The model is evaluated on a large EHR dataset of 19,218 patients and 368,790 visits. (iv) It achieves high performance with balanced sensitivity and specificity. The framework outperforms convolutional neural network–long short-term memory (CNN-LSTM), bidirectional long short-term memory (Bi-LSTM), LSTM, support vector machine (SVM), and k-nearest neighbor (KNN) baseline models. (v) This approach provides a scalable and clinically relevant solution for early diabetes risk prediction.

## II. Related Work

Early work on EHR-based diabetes risk modeling established the foundation for population-level prediction. Hippisley-Cox *et al*. [7] developed QDiabetes to estimate 10-year risk via routinely collected primary-care data. This model offered a scalable approach to population stratification but was limited by its cross-sectional structure, which does not account for dynamic changes within individuals over time. Carrasco-Ribelles *et al*. [8] reviewed longitudinal prediction models and reported that sequence-aware approaches outperform static baselines when they capture visit order, spacing, and persistence of clinical signals. Their review also revealed heterogeneous preprocessing strategies, inconsistent prediction windows, and poor calibration reporting, which complicate study comparison and clinical translation. Petridis *et al*. [12] highlighted the frequent reliance on narrow feature sets and coarse temporal representations, whereas Khokhar *et al*. [13] noted improved performance metrics but emphasized the lack of transparent code, standardized validation protocols, and decision curve evaluations.

Temporal modeling has demonstrated clear advantages. Swinckels *et al*. [10] reported that recurrent and attention-based models improve predictive performance by utilizing full visit sequences rather than aggregated snapshots. Despite these gains, many studies compressed laboratory and vital measurements into broad intervals, losing the temporal precision critical for identifying early risk transitions. Liu *et al*. [9] further demonstrated the strength of longitudinal modeling via decade-long primary care records but did not validate their models across multiple health systems, limiting generalizability.

Hybrid architectures were introduced to integrate hierarchical feature representations with temporal memory. Tian *et al*. [14] used convolutional encoders with LSTM layers to enhance EHR event prediction, achieving superior performance compared with single-stage models. Krishnankutty [15] reported similar improvements in readmission risk prediction. A common limitation remains evident across these approaches: intermediate convolutional representations are often discarded, with only the terminal feature map propagated to the recurrent layer. This practice risks losing informative patterns that emerge at earlier stages of the network.

Feature engineering grounded in pathophysiology has also shown substantial predictive value but remains underutilized. Yuge *et al*. [11] demonstrated that the triglyceride-to-HDL-cholesterol ratio strongly predicts incident T2D and often outperforms individual lipid components. Zhang *et al*. [16] and Tan *et al*. [17] reported significant associations between total cholesterol/HDL and non-HDL/HDL ratios and future T2D in large population cohorts. These compact biomarkers provide interpretable indicators of insulin resistance and cardiometabolic stress. However, most temporal models rely on raw lipid values or BMI alone and do not incorporate these clinically validated composite markers into sequence representations.

Consequently, several methodological gaps persist. Prior work frequently (i) aggregates time series data into coarse intervals. This weakens the ability to capture short- and mid-range dynamics [8-10]; (ii) It discards intermediate convolutional features in hybrid CNN–RNN architectures. This reduces representational depth [14, 15]; (iii) it underutilizes composite biomarkers such as TG/HDL, TC/HDL, non-HDL/HDL, and adiposity indicators [11, 17]; and (iv) it inconsistently reports calibration and clinical utility. This limits real-world applicability [8, 13]. This study addresses these gaps. It preserves multilevel convolutional features, routes each to a dedicated recurrent module, and integrates clinically meaningful lipid ratios and adiposity predictors to improve early T2D risk estimation.

Statement of Significance: (i) Problem or issue: Type 2 diabetes constitutes a major global health burden associated with high morbidity, mortality, and economic cost. Early identification of individuals at elevated risk is crucial for implementing preventive strategies. Existing predictive models often rely on static representations of electronic health record (EHR) data. They fail to capture the temporal evolution of clinical variables that precede disease onset; (ii) what is already: Conventional machine learning and deep learning frameworks have demonstrated improved predictive performance compared with traditional statistical models. However, most current approaches employ single-level temporal features and limited clinical inputs. They do not adequately represent short- and intermediate-term variations that characterize the transition from prediabetes to diabetes. Clinically relevant biomarkers such as lipid ratios and obesity indices are rarely incorporated, which limits model interpretability and clinical translation; (iii) what this paper adds: This study proposes a hybrid deep learning framework that integrates hierarchical temporal modeling with clinically engineered biomarkers. The model extracts temporal features at multiple abstraction levels and incorporates validated predictors such as TG/HDL, LDL/HDL, TC/HDL, VLDL, and obesity status. Compared with the CNN-LSTM, Bi-LSTM, SVM, and KNN baselines, it achieved 93.2% accuracy and demonstrated superior sensitivity and specificity. The proposed architecture enhances interpretability, robustness, and scalability, providing a clinically aligned approach to early diabetes risk prediction; (iv) who would benefit clinicians and healthcare institutions, who can employ this framework for timely identification of high-risk individuals and for guiding personalized preventive interventions. Researchers may adapt the model for other chronic conditions characterized by longitudinal clinical data. Health policy makers and digital health developers can leverage the methodology to strengthen predictive analytics and optimize population-level disease management strategies.

## III. Methods

The proposed methodological framework, illustrated in Fig. 1, is organized into six sequential phases. The process begins with dataset acquisition and data preprocessing, which ensure data reliability and proper temporal alignment across patient records. Through feature engineering, clinically meaningful and derived biomarkers are integrated to enrich the predictive space. Hierarchical temporal extraction applies multiscale convolutional pooling to capture low-, mid-, and high-level temporal patterns. The temporal modeling phase employs parallel LSTM modules to encode sequential dependencies and generate a unified temporal representation. The Model Development stage integrates these fused features within a hybrid CNN–LSTM framework to predict diabetes risk. Finally, evaluation metrics such as accuracy, sensitivity, specificity, precision, F1 score, and the Matthews correlation coefficient are used to quantify predictive performance. This structured framework establishes a comprehensive approach for temporal learning and clinically interpretable disease risk prediction.

**Fig. 1.**
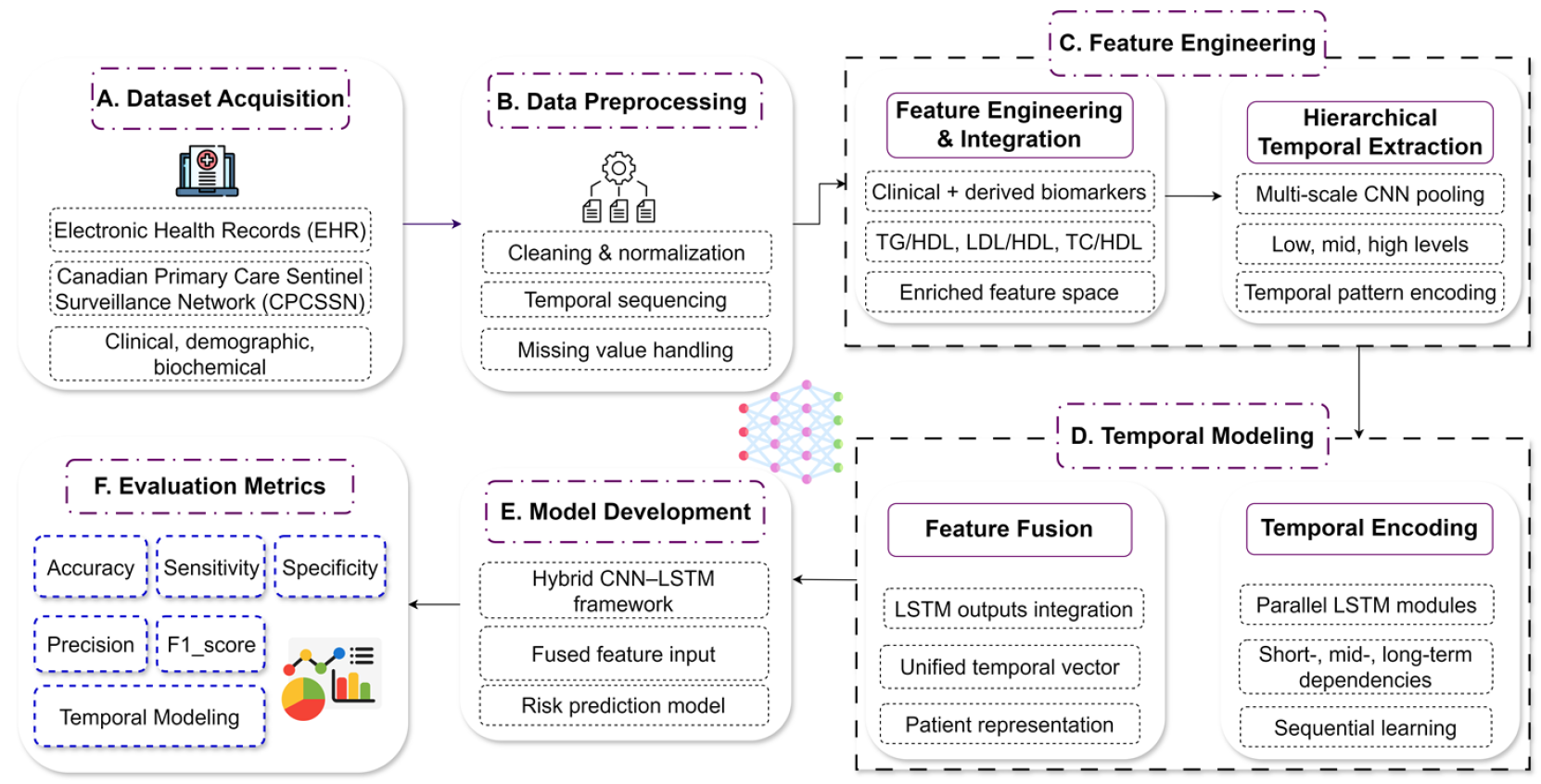
Architecture of the proposed CNN–LSTM–based temporal modeling framework. It includes six phases: dataset acquisition, data preprocessing, feature engineering, temporal modeling, model development, and evaluation metrics for diabetes risk prediction.

### A. Dataset Acquisition

The dataset was obtained from the Canadian Primary Care Sentinel Surveillance Network (CPCSSN) [18]. It spans the years 1998–2015 and includes 19,218 adult patients aged 18–90 years, with a total of 368,790 longitudinal visit records. The cohort consisted of 11,054 women and 8,164 men. At the end of follow-up, 7,719 individuals met the diagnostic criteria for type 2 diabetes, whereas 11,499 remained nondiabetic. At least two recorded visits and complete demographic identifiers were required for inclusion. Patients who were diagnosed with diabetes at their first recorded visit were excluded. For incident diabetes cases, records were retained up to the initial diagnosis. For nondiabetic controls, all visits except the final visit were used to enable next-visit risk prediction. The dataset represents a diverse primary care population with detailed longitudinal data. Figure 2 presents the distribution of patients by diabetic status and sex, emphasizing the cohort’s demographic balance. The main clinical and demographic characteristics are summarized in Table 1, which details the anthropometric, biochemical, and comorbidity profiles essential for predictive modeling. Each patient record includes 14 core variables. These variables include demographic information (patient ID, age, sex), clinical observations (BMI, systolic blood pressure), laboratory measurements (FBS, A1c, HDL, LDL, TG, total cholesterol), and clinical diagnoses (hypertension, osteoarthritis, COPD, depression).

**Table 1.** Baseline characteristics of the study dataset. The table summarizes demographic information, clinical observations, laboratory measurements, and clinical diagnoses of the study population.

| Demographic Information | Findings |
| --- | --- |
| Samples | 368790 |
| Patients | 19218 |
| Female patients, N (%) | 11054 (57.5) |
| Male patients, N (%) | 8164 (42.4) |
| Age, mean ± SD, Years | 65 ± 12.5 |
| **Clinical Observation** |  |
| sBP, mean ± SD, mm Hg | 132.05 ± 17.1 |
| BMI, mean ± SD, kg/m<sup>2</sup> | 30.71 ± 9.64 |
| **Lab Values** |  |
| FBS, mean ± SD, mmol/L | 6.11 ± 1.58 |
| A1C, mean ± SD, mmol/L | 6.4 ± 0.93 |
| TG, mean ± SD, mmol/L | 1.63 ± 1.01 |
| HDL, mean ± SD, mmol/L | 1.35 ± 0.4 |
| LDL, mean ± SD, mmol/L | 2.7 ± 1.05 |
| Cholesterol mean ± SD, mmol/L | 4.79 ± 1.23 |
| BMI, mean ± SD, kg/m<sup>2</sup> | 30.71 ± 9.64 |
| **Clinical Diagnosis** |  |
| Hypertension, N (%) | 13766 (71.6) |
| COPD, N (%) | 2138 (11.1) |
| OA, N (%) | 7274 (37.8) |
| Depression, N (%) | 4512 (23.4) |

**Fig. 2.**
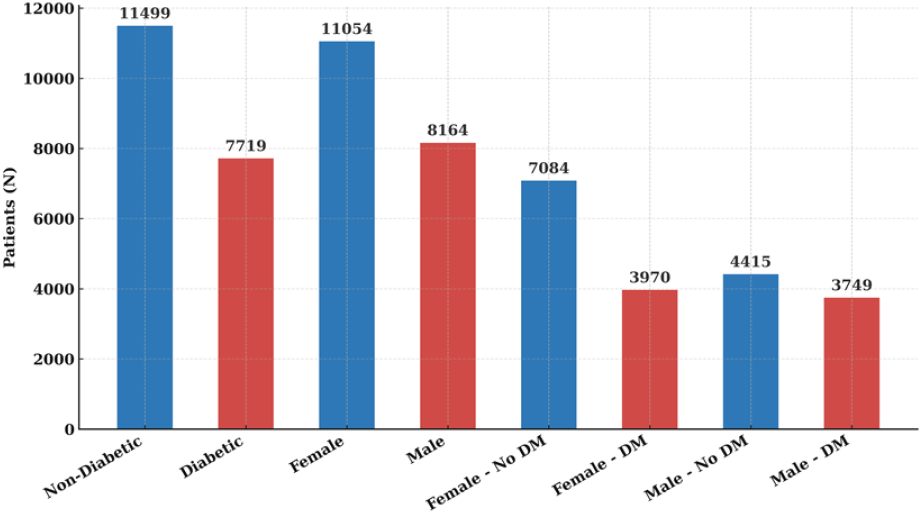
Distribution of patients across diabetic and non-diabetic groups by sex. The figure presents the proportions of male and female participants in each category.

### B. Data Preprocessing

This stage involved cleaning and normalization, temporal sequencing, and missing value handling. Data cleaning removed duplicate entries and inconsistent records. Clinical and laboratory variables were normalized to eliminate scale-related bias and ensure comparability across patients. Patient visits were then arranged in chronological order to preserve the temporal context of disease progression. Each patient sequence represented a continuous timeline of clinical observations and laboratory results. Missing values within individual sequences were imputed using the cohort mean [19]. The outliers were replaced with the most recent valid value from the same patient on the basis of the last observation carried forward (LOCF) approach [20]. For incident diabetes patients, sequences were retained up to the first diagnosis. For nondiabetic controls, all visits except the final visit were included to support next-visit prediction. After preprocessing, the data were transformed into fixed-length feature vectors. The dataset was divided into training (80%) and testing (20%) subsets via stratified sampling to maintain class balance.

### C. Feature Engineering

Feature engineering aims to strengthen the model’s predictive capacity and represent clinically significant biochemical and temporal patterns. The process included two main stages: feature engineering and integration and hierarchical temporal extraction. In the first stage, derived biochemical indicators were incorporated to extend the dataset beyond standard clinical variables. On the basis of established metabolic associations, six engineered predictors have been introduced: prediabetic status, TG/HDL, LDL/HDL, TC/HDL, VLDL, and obesity status [21, 22]. As summarized in Table 2, these variables combined continuous and binary attributes obtained from laboratory and clinical assessments. Prediabetic status was defined as fasting blood glucose between 5.5 and 6.9 mmol/L, according to standard diagnostic guidelines [23]. The TG/HDL ratio serves as an indicator of insulin resistance and cardiovascular risk [24, 25]. VLDL was estimated by dividing triglycerides by 2.2. Ratios such as LDL/HDL and TC/HDL reflect lipid imbalances associated with metabolic syndrome and early diabetes onset [26, 27]. Obesity was determined via body mass index (BMI), a well-recognized metabolic risk indicator [28]. All continuous features were standardized, and categorical features were binarized to maintain a uniform data structure. These enriched attributes were integrated with demographic and clinical variables to produce an expanded feature space for model training.

**Table 2.** Engineered diabetic predictors derived from clinical and biochemical variables. The table lists each attribute, its measurement unit, description, and feature type used for model development.

| Attribute | Units | Description | Feature Type |
| --- | --- | --- | --- |
| Pre-diabetic | N% | Lab Result | Binary |
| TG/HDL | mmol/L | Lab Result | Continuous |
| LDL/HDL | mmol/L | Lab Result | Continuous |
| VLDL | mmol/L | Lab Results | Continuous |
| Total Cholesterol/HDL | mmol/L | Lab Results | Continuous |
| Obese | N% | Clinical Observation | Binary |

Feature relevance was assessed through two complementary analytical methods: classification and regression tree (CART)-based feature importance and univariate chi-square analysis [29, 30]. The CART algorithm quantifies the contribution of each predictor to model decisions and assigns an importance score proportional to its discriminative strength. As shown in Fig. 3(a), the most influential predictors were fasting blood sugar (FBS), prediabetic status, and glycated hemoglobin (A1c). Lipid ratios such as TG/HDL, VLDL, and TC/HDL followed in predictive importance. High-density lipoprotein (HDL) has lower scores, which is consistent with its protective metabolic function. The chi-square test in Fig. 3(b) supported these findings. The analysis validated the same variables as statistically significant discriminators of diabetes risk.

**Fig. 3.**
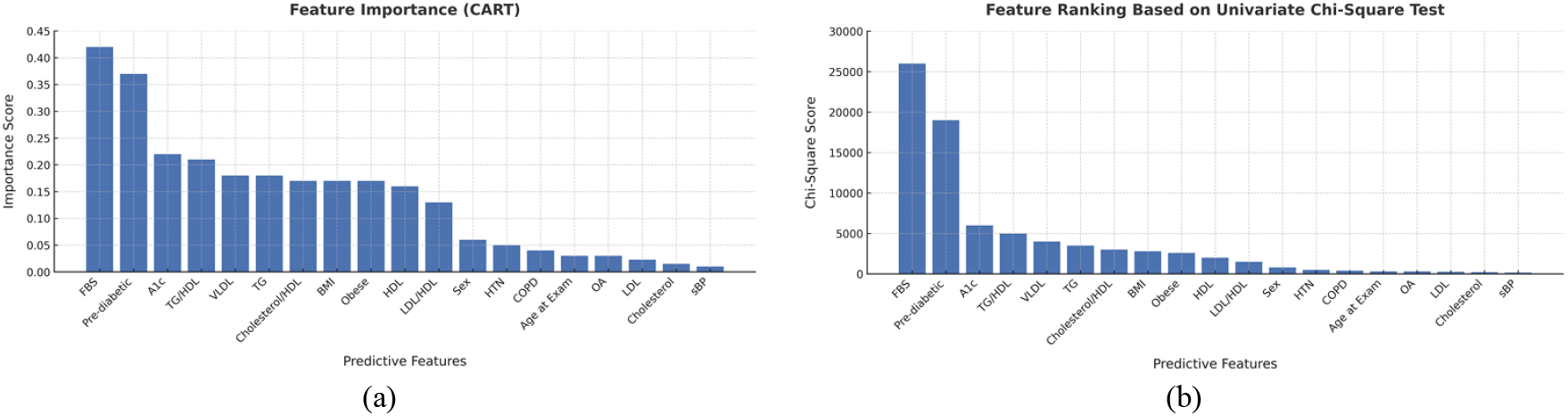
Feature relevance evaluation using statistical and tree-based methods. (a) Feature importance scores derived from the Classification and Regression Tree (CART) method demonstrate the relative contribution of predictive variables to diabetes risk. (b) Univariate chi-square test identifies the most statistically significant categorical and continuous predictors associated with diabetes.

### D. Temporal Modeling

Temporal modeling aims to capture dynamic dependencies across patient visit sequences to improve disease risk prediction. This stage consists of two main processes, feature fusion and temporal encoding, as illustrated in the methodological framework. In the feature fusion step, feature representations from multiple convolutional and recurrent layers are integrated to form a unified temporal feature vector. Outputs from the LSTM layers were aggregated to consolidate multilevel information extracted during earlier feature learning. This integration enabled the construction of a single temporal vector representing each patient’s longitudinal health trajectory. The fused representation provided a comprehensive summary of short- and long-term visit dependencies, forming a robust input for subsequent prediction layers. The temporal encoding step uses parallel LSTM modules to model sequential patterns and capture dependencies at various time scales—short, intermediate, and long-term. Each module independently processed feature sequences derived from different CNN pooling levels, ensuring that both rapid clinical changes and gradual disease progression were effectively represented. The outputs of these parallel modules were then concatenated to form a rich temporal encoding that emphasized the progression of metabolic and cardiovascular risk factors over time. Through sequential learning, the model preserved contextual continuity between consecutive visits, allowing the system to infer patterns associated with the onset and progression of type 2 diabetes. This hierarchical recurrent design ensures a balanced representation of both local fluctuations and long-term clinical evolution, which are critical for accurate temporal risk assessment.

### E. Model Development

The proposed framework was developed as a hybrid CNN–LSTM architecture to capture spatial and temporal patterns jointly within patient visit sequences. It consists of two primary components: feature extraction and risk prediction. In the feature extraction stage, the CNN component automatically learns hierarchical temporal representations from sequential health records. The multilevel pooling layers extract feature vectors from low-, mid-, and high-level abstractions, denoted as *F* = [*F*_1_,*F*_2_,…,*F*_*n*_]^*L*^, where *L* represents the layer depth. Unlike conventional single-scale feature extraction, this design preserves hierarchical information by maintaining distinct representations across pooling layers. The extracted vectors are then passed to independent LSTM layers to model temporal dependencies within each level.

Let 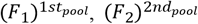, and 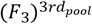 represent the feature sets derived from successive pooling stages. These are processed by their corresponding LSTM blocks, allowing each to specialize in capturing short-, mid-, or long-term sequential dynamics. The outputs from all the LSTM blocks are concatenated to form a fused feature input, producing a unified temporal vector that represents each patient’s longitudinal profile. The concatenated output is computed via Eq. (1):

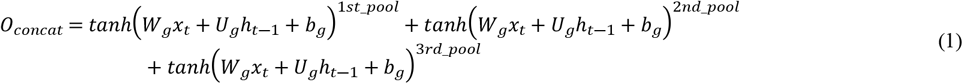

where *O*_*concat*_ is the concatenated output from all the LSTM layers, *tanh* is the activation function, *W*_*g*_ and *U*_*g*_ are weight matrices, and *b*_*g*_ is the bias vector. In the risk prediction stage, the fused temporal vector is passed through fully connected dense layers to estimate the probability of diabetes onset. Dropout and global max pooling were applied to reduce overfitting and improve generalizability. This hybrid CNN–LSTM framework effectively integrates spatial abstraction with temporal learning, allowing the model to recognize both rapid fluctuations and gradual changes in clinical indicators across time. By preserving multilevel temporal representations, the model captures complex clinical dependencies and enhances interpretability in predicting diabetes progression.

### F. Evaluation Metrics

To evaluate the performance of the proposed hybrid CNN–LSTM model, a comprehensive set of established performance metrics was applied. These metrics include accuracy, sensitivity, specificity, precision, F1 score, and the Matthews correlation coefficient (MCC) [31]. Each metric captures a different perspective of predictive performance. Using multiple metrics ensures a comprehensive and balanced assessment. Accuracy represents the overall proportion of correct predictions. Sensitivity (recall) measures the ability to identify positive cases. Specificity reflects how well the model recognizes negative cases. Precision evaluates the proportion of predicted positives that are truly positive. The F1 score combines sensitivity and precision into a single measure. It is particularly useful when the class distribution is imbalanced. The MCC incorporates all four outcomes: true positives, true negatives, false positives, and false negatives. It provides a more robust evaluation of binary classification performance, even with skewed datasets. The mathematical definitions of these metrics are presented in Eqs. (2) – (7):

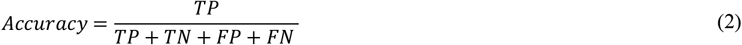

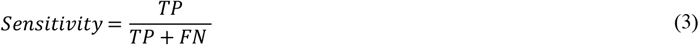

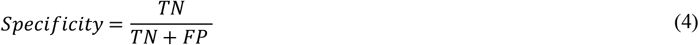

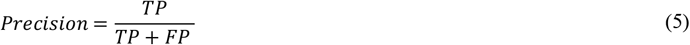

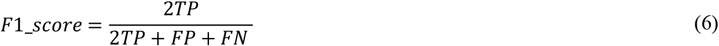

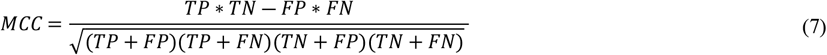

where TP refers to true positives, TN refers to true negatives, FP refers to false positives, and FN refers to false negatives. The MCC ranges from −1 to +1. A value of +1 indicates a perfect prediction. A value of 0 corresponds to random prediction. A value of −1 indicates complete disagreement between the predictions and the ground truth.

## IV. Results

### A. Experimental setup

The experimental protocol was designed to ensure reproducibility, computational efficiency, and fair comparison across baseline models. All the experiments were executed on a workstation equipped with an Intel Core i7-10750H processor (2.60 GHz), 16 GB of RAM, and an NVIDIA GeForce RTX 1080 GPU, which provided stable performance for both the training and evaluation phases. To establish a rigorous performance benchmark, the proposed model was evaluated alongside five widely used methods: KNN, SVM, LSTM, CNN-LSTM, and Bi-LSTM. Two independent training cohorts were randomly sampled from the original dataset, comprising 3,892 and 6,281 patients. This dual-cohort design enabled a systematic assessment of the impact of the data scale and feature representation on model performance. Each model was trained under two feature configurations: (1) the original clinical variables and (2) an integrated feature set that combined original variables with engineered predictors derived from lipid ratios, prediabetic indicators, and obesity status. This factorial design resulted in a total of 24 experiments, covering variations in architecture, feature configuration, and cohort size. The proposed framework integrates multilevel CNN feature extraction, parallel LSTM temporal encoding, and a late fusion strategy, allowing it to learn both localized patterns and long-range temporal dependencies.

### B. Model Performance

The proposed architecture demonstrated a clear and consistent performance advantage across all the experimental settings. It achieved the highest accuracy among all the evaluated methods, regardless of dataset size or feature configuration. For the larger cohort of 6,281 patients, the model attained 93.2% accuracy with the integrated feature set and 92.2% accuracy with the original set. When trained on the smaller cohort of 3,892 patients, the accuracy remained strong at 90.5% and 89.8%, respectively. This stability indicates robust generalizability and reliable predictive capacity even when data availability is limited. This consistent superiority becomes evident when its performance is compared with that of both classical and temporal baselines. Traditional classifiers such as K-nearest neighbors (KNNs) and support vector machines (SVMs) struggle to capture temporal dependencies in clinical data, resulting in lower accuracy. Temporal deep learning models, including CNN-LSTM and Bi-LSTM, demonstrated improved results but were still outperformed by the proposed multilevel fusion model, highlighting the added value of their hierarchical temporal representation. These trends are quantitatively supported by the results presented in Table 3. The performance margin remains clear at both data scales, reflecting the model’s ability to exploit both short-term fluctuations and long-term disease patterns embedded in longitudinal records. In addition to overall accuracy, Table 4 reveals the strength of the model across key diagnostic metrics. For the 6,281-patient cohort with integrated features, 75.7% sensitivity, 98.8% specificity, 95.3% precision, 84.4% F1 score, and 81.0% MCC were achieved. This balance between sensitivity and specificity is particularly critical in clinical risk stratification, where false negatives and false positives have different but equally important consequences. The model’s ability to maintain this equilibrium underscores its potential suitability for real-world decision support systems in early diabetes risk prediction.

**Table 3.** Performance comparison of the proposed hybrid model with state-of-the-art machine learning and deep learning architectures. The table presents accuracy across varying data densities, using both original and integrated feature sets.

| Model | Training Data Density (%) | Original Feature (%) | Integrated Feature Set (%) |
| --- | --- | --- | --- |
| KNN | 3892 | 54.0 | 80.5 |
|  | 6281 | 80.0 | 89.3 |
| SVM | 3892 | 82.9 | 88.8 |
|  | 6281 | 88.9 | 90.8 |
| LSTM | 3892 | 85.8 | 87.8 |
|  | 6281 | 89.1 | 90.9 |
| Bi-LSTM | 3892 | 86.0 | 88.0 |
|  | 6281 | 89.6 | 90.8 |
| CNN-LSTM | 3892 | 86.3 | 88.5 |
|  | 6281 | 90.1 | 91.1 |
| Proposed Model | 3892 | 89.8 | 90.5 |
|  | 6281 | 92.2 | 93.2 |

**Table 4.** Performance metrics of the proposed hybrid model compared with benchmark algorithms. Results include accuracy, sensitivity, specificity, precision, F1 score, and Matthews correlation coefficient (MCC) using both original and merged feature sets.

| Model | Feature Set | Accuracy (%) | Sensitivity (%) | Specificity (%) | Precision (%) | F1 Score (%) | MCC |
| --- | --- | --- | --- | --- | --- | --- | --- |
| Proposed Model | Original | 92.2 | 75.7 | 97.7 | 91.4 | 82.8 | 78.5 |
|  | Merged | **93.2** | 75.7 | **98.8** | **95.3** | **84.4** | **81.0** |
| LSTM | Original | 89.1 | 71.5 | 94.7 | 81.1 | 76.0 | 69.2 |
|  | Merged | 90.9 | 68.3 | 98.1 | 92.1 | 78.4 | 74.2 |
| Bi-LSTM | Original | 89.6 | 72.2 | 95.1 | 82.5 | 77.0 | 70.6 |
|  | Merged | 90.8 | 74.9 | 95.9 | 85.5 | 79.8 | 74.3 |
| CNN-LSTM | Original | 90.1 | 64.1 | 98.4 | 92.7 | 75.8 | 71.7 |
|  | Merged | 91.0 | 71.2 | 97.3 | 89.4 | 79.2 | 74.4 |
| SVM | Original | 88.9 | 73.6 | 93.7 | 79.0 | 76.2 | 69.0 |
|  | Merged | 90.8 | 74.9 | 95.8 | 85.2 | 79.7 | 74.1 |
| KNN | Original | 80.0 | **82.3** | 79.2 | 55.8 | 66.5 | 55.0 |
|  | Merged | 89.3 | 60.1 | 98.6 | 93.4 | 73.1 | 69.4 |

### C. Feature contribution and data-scale effects

The results highlight the critical impact of clinically informed feature engineering on predictive accuracy. Integrating prediabetic indicators, lipid ratios, and obesity markers expanded the input space and improved model discriminability across all evaluated deep learning architectures. The proposed model demonstrated the greatest performance gain, particularly in the larger patient cohort. This trend is illustrated in Fig. 4(a) for 6,281 patients and Fig. 4(b) for 3,892 patients. The training data scale was another decisive factor influencing performance. Increasing the cohort from 3,892 to 6,281 patients improved the accuracy of every evaluated model, with the largest absolute gains observed in the proposed architecture. This improvement reflects the model’s capacity to learn more complex temporal representations when exposed to a broader and more diverse patient population. Compared with deep temporal models, classical classifiers achieve minimal gains with scale expansion, further confirming their limited adaptability. These outcomes can be attributed to the multilevel temporal encoding design. The architecture captures early, intermediate, and late convolutional features, preserving both local fluctuations and long-range temporal dependencies. The separate LSTM modules process these features independently, while the late fusion layer consolidates them into a unified representation. This structured encoding allows the model to detect subtle temporal signals and capture disease progression dynamics more effectively than single-stream temporal approaches do. As a result, the system delivers not only higher accuracy but also more balanced diagnostic performance, providing a solid foundation for scalable and clinically meaningful predictive modeling.

**Fig. 4.**
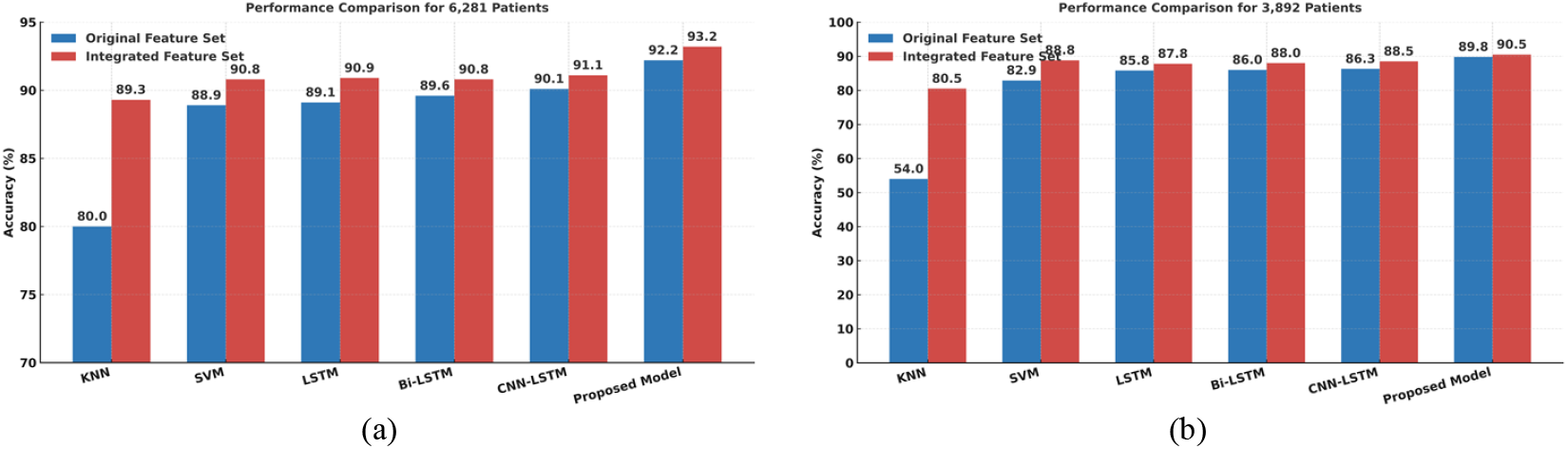
Model accuracy across original and integrated feature sets. (a) Results for 6,281 patients showing higher accuracy with the integrated features. (b) Results for 3,892 patients confirming consistent performance gains across all models.

## V. Discussion

### A. Principal Results

Diabetes is a major global health problem. It causes significant morbidity, mortality, and economic strain on healthcare systems. In 2024, diabetes accounted for more than 3.4 million deaths worldwide [3]. Early prediction enables timely intervention and targeted prevention. This study addresses this need with a temporal deep learning framework that provides stable and accurate risk prediction. The proposed multilevel temporal model achieved an accuracy of 93.2% on the larger cohort of 6,281 patients and 90.5% on the smaller cohort of 3,892 patients. The competing models showed a marked decline when the dataset size decreased. The stable performance of the proposed framework reflects its ability to generalize temporal patterns in clinical records. The analysis used the CPCSSN dataset, which includes 368,790 unique patient records collected between 1998 and 2015. This large longitudinal resource supports realistic evaluation under complex clinical data conditions.

Feature engineering contributed substantially to the observed performance. The model incorporated prediabetic indicators [32], lipid ratios (TG/HDL, LDL/HDL, TC/HDL, and VLDL), and obesity markers. Models trained only on the original variables showed limited predictive capacity. Adding these engineered features increased the sensitivity to 75.7% and the MCC to 81.0%. These predictors reflect well-established links to insulin resistance, β-cell dysfunction, and metabolic shifts before disease onset [33, 34]. The temporal architecture captures patterns at multiple scales. Early convolutional layers detect short-term fluctuations, intermediate layers extract transitional trends, and late layers retain long-range dependencies. Each temporal representation passes through a separate LSTM stream, and a fusion layer merges the outputs into a single decision signal. This design preserves temporal information and maintains high diagnostic performance, with a specificity of 98.8% and a sensitivity of 75.7%. Feature importance analysis confirmed that fasting blood sugar, HbA1c, and prediabetic status were the strongest predictors. This combination of temporal encoding and clinical features delivers sharper decision boundaries, stronger generalization, and stable performance across cohorts. The framework functions as a clinically meaningful predictive system rather than a narrowly tuned algorithm.

### B. Comparative Performance

The proposed architecture outperformed all the baseline models across all the settings. With 6,281 patients and integrated features, the accuracy reached 93.2%. The best temporal baseline was CNN-LSTM [35] at 91.1%, followed by Bi-LSTM [54] at 90.8% and LSTM [36] at 90.9%. The classical models trailed further behind, including SVM [37] at 90.8% and KNN [38] at 89.3%. In the smaller cohort of 3,892 patients, the ranking remained unchanged. The proposed model achieved 90.5%, whereas the CNN-LSTM reached 88.5%, the Bi-LSTM reached 88.0%, the LSTM reached 87.8%, the SVM reached 88.8%, and the KNN reached 80.5%. This consistent lead demonstrates stable performance across different scales and configurations. Accuracy alone did not explain this advantage. In the 6281-patient cohort, the model achieved a sensitivity of 75.7%, specificity of 98.8%, precision of 95.3%, F1 score of 84.4%, and MCC of 81.0% [38, 39]. Competing temporal models showed lower sensitivity and MCCs at similar specificities. The classical methods either sacrifice sensitivity for specificity or increase the number of false positives to increase recall. The proposed model maintained a more balanced diagnostic profile, which is critical for clinical risk stratification. Feature integration increased the performance gap. Replacing the original variables with the integrated feature set improved the accuracy for all the models. The largest absolute gains appeared in the proposed model and CNN-LSTM, indicating that clinically engineered predictors, prediabetes status, lipid ratios (TG/HDL, LDL/HDL, TC/HDL, and VLDL), and obesity carry complementary signals [23, 28]. The proposed model leveraged this signal most effectively through its multilevel temporal pathways and late fusion strategy. It also demonstrated better scalability. When the cohort size increased from 3,892 to 6,281 patients, the accuracy improved across all approaches, but the proposed model preserved the largest lead and showed the smallest relative decline at lower scales. This behavior reflects its architecture, where early, intermediate, and late convolutional features feed separate LSTM streams before fusion. This design preserves both short-horizon fluctuations and long-range trends more effectively than single-stream encoders do. Overall, the model achieved higher accuracy, better sensitivity–specificity balance, and stronger scale resilience than state-of-the-art temporal baselines (LSTM, Bi-LSTM, and CNN-LSTM) and classical comparators (SVM and KNN). The performance gains stem from hierarchical temporal representation combined with clinically grounded feature integration, not from model depth alone.

### C. Architectural and Feature Engineering Innovations

The model applies multilevel temporal encoding to capture disease dynamics at different time scales. Low-, mid-, and high-level convolutional feature routes to parallel LSTM streams that represent short, transitional, and long temporal patterns. A late-fusion layer merges these outputs into a single decision vector. This hierarchical structure preserves temporal precision and improves abstraction in longitudinal clinical data [40, 41]. The convolutional backbone functions as a multiresolution extractor. Early layers highlight rapid fluctuations. Intermediate layers reflect evolving physiological trends. The deep layers encode stable disease signatures. This design provides richer evidence than single-stream CNN-LSTM or Bi-LSTM pipelines do [42, 43]. Fusion uses a gated mechanism to weight temporal streams on the basis of their relevance at the decision layer, which improves the signal-to-noise ratio compared with simple concatenation [40, 44, 45].

The model integrates clinically engineered biomarkers with temporal features. These biomarkers include prediabetes status, TG/HDL, LDL/HDL, TC/HDL, VLDL, and obesity. They are strongly linked to insulin resistance, β-cell dysfunction, and early metabolic shifts. Their inclusion enhances the model’s ability to handle class imbalance and irregular sampling. Feature importance analysis confirmed that fasting blood sugar, HbA1c, and prediabetic status were the most discriminative predictors. These features anchor the framework to clinically validated risk signals. Dropout and global max pooling stabilize the network while preserving feature richness. The design maintains precision and recall across different cohort sizes [43, 46]. Together, hierarchical temporal routing and clinically grounded biomarkers form the core innovation. The framework advances temporal EHR modeling beyond terminal-map CNN-LSTM baselines.

### D. Clinical Impact

The proposed framework offers direct clinical value for early diabetes risk identification and preventive management. Its multilevel temporal structure detects subtle disease trajectories before overt onset. Capturing both short-term fluctuations and long-range metabolic trends enables timely risk stratification at the individual level [46]. This early detection supports proactive interventions that can delay or prevent the progression from prediabetes to diabetes [47]. The integration of clinically engineered predictors strengthens this utility. Lipid ratios and prediabetes indicators are well-established metabolic markers with strong predictive value [45, 48, 49]. Embedding these features within the temporal model aligns algorithmic outputs with clinically interpretable signals. This reduces the gap between predictive models and routine decision making. It allows clinicians to link high-risk alerts to specific physiological markers rather than latent representations [50, 51]. The model’s stability across data scales ensures effective use in both large hospitals and small community clinics [52]. This scalability supports integration with existing EHR infrastructures. Temporal dynamics are preserved, enabling risk profiles to update at each patient visit without retraining. This supports continuous learning health systems and aligns with personalized and precision medicine [53].

From a clinical workflow perspective, the framework functions as a decision support layer. Its outputs can be fed into clinical dashboards to flag high-risk patients for follow-up testing or lifestyle interventions. This targeted approach is more cost-effective than population-wide screening is [54]. The engineered predictors provide explainable anchors that increase clinician trust and support adoption. The framework also has translational potential beyond diabetes. Hierarchical temporal encoding and clinically grounded features are adaptable to other metabolic and cardiovascular conditions. Early detection and longitudinal tracking remain central to these diseases [55, 56]. This architecture establishes a foundation for robust, scalable, and interpretable predictive systems across diverse clinical settings.

### E. Limitations and Future Work

Despite its strong performance, the proposed framework has several limitations. The model was trained and validated within the Canadian Primary Care Sentinel Surveillance Network, which is national in scope and includes approximately 1.2 million patients from approximately 1,100 primary care practices across eight provinces. The results therefore reflect Canadian primary care only. Transportability to other EHR networks, to hospital or specialty settings, and to health systems outside Canada remains untested. Site and system differences in coding practices, missingness patterns, and population characteristics can affect performance [40]. External validation on independent multi-institutional datasets, including non-CPCSSN and cross-national cohorts, is needed to assess robustness [57]. The dataset also lacked variables such as family history of diabetes, polycystic ovary syndrome, and skinfold thickness, which are established risk markers [58, 59]. Unstructured data such as clinical notes and radiology reports were not used. Adding these elements through natural language processing may improve coverage and context.

The temporal architecture assumes a uniform data structure after preprocessing. Real-world EHR data often contain irregular visit intervals and incomplete records [40]. Advanced designs, such as time-aware attention or continuous-time neural models, can address these challenges [60, 61]. Parts of the model, including the fusion and recurrent layers, remain partially opaque. Applying explainability methods such as attention weight visualization, concept-based attribution, or counterfactual reasoning may increase clinical trust [62]. The framework has not yet been evaluated prospectively. Retrospective analysis does not capture operational issues such as data latency, integration with clinical workflows, or physician response. Prospective clinical studies are needed to assess real-world performance and usability. Finally, the framework can be extended to other chronic conditions. Hierarchical temporal encoding and clinically engineered predictors are adaptable to cardiovascular and metabolic diseases. Federated learning may further support scalability while preserving data privacy across institutions [63].

## VI. Conclusion

This study presents a hybrid deep learning framework that combines hierarchical temporal modeling with clinically engineered biomarkers for early diabetes risk prediction. The framework uses convolutional neural network (CNN) feature extraction and parallel long short-term memory (LSTM) modules. It captures short-term fluctuations, intermediate transitions, and long-term trends in longitudinal electronic health record (EHR) data. The model includes clinically meaningful indicators. These include the triglyceride-to-high-density lipoprotein cholesterol ratio (TG/HDL-C), low-density lipoprotein to high-density lipoprotein cholesterol ratio (LDL/HDL-C), total cholesterol to high-density lipoprotein cholesterol ratio (TC/HDL-C), very low-density lipoprotein (VLDL), prediabetes status, and obesity. These biomarkers provide a physiological basis that improves model interpretability and reliability. The proposed framework achieved 93.2% accuracy with balanced sensitivity and specificity. It outperformed classical machine learning algorithms and standard temporal deep learning models. Its stable performance across different cohort sizes confirms its scalability and suitability for clinical settings. The use of validated biomarkers aligns the model with existing clinical practices. These findings support early identification of high-risk individuals and enable targeted preventive interventions. This can reduce the progression of diabetes and decrease the economic burden on healthcare systems. Future work will expand the model to multicenter datasets. Sociodemographic and lifestyle variables will be included. In addition, it incorporates explainable artificial intelligence. The architecture can also support risk prediction for other chronic diseases. This position positions the framework as a scalable and clinically relevant tool for predictive healthcare.

## Data Availability

The dataset analyzed in this study was accessed through the Canadian Primary Care Sentinel Surveillance Network (CPCSSN) under a formal data-sharing agreement. Due to privacy and confidentiality regulations, the data cannot be made publicly available. Researchers who meet the criteria for access to sensitive health data may obtain it directly from CPCSSN at https://www.cpcssn.ca.

## Author Contributions

**Conceptualization:** Iqra Naveed, Muhammad Farhat Kaleem, Karim Keshavjee, Aziz Guergachi.

**Data curation:** Iqra Naveed, Mohammad Noaeen, Karim Keshavjee.

**Formal analysis:** Iqra Naveed.

**Investigation:** Iqra Naveed, Muhammad Farhat Kaleem, Aziz Guergachi.

**Methodology:** Iqra Naveed, Muhammad Farhat Kaleem.

**Project administration:** Muhammad Farhat Kaleem.

**Supervision:** Karim Keshavjee.

**Validation:** Karim Keshavjee.

**Visualization:** Iqra Naveed, Mohammad Noaeen.

**Writing – original draft:** Iqra Naveed, Mohammad Noaeen.

**Writing – review & editing:** Muhammad Farhat Kaleem, Mohammed A. AboArab, Karim Keshavjee, Aziz Guergachi.

**Declaration of Competing Interest**

The authors declare no known competing financial interests or personal relationships that could have influenced the work reported in this paper.

